# Even one metre seems generous. A reanalysis of data in: Chu *et al*. (2020) Physical distancing, face masks, and eye protection to prevent person-to-person transmission of SARS-CoV-2 and COVID-19

**DOI:** 10.1101/2020.06.11.20127415

**Authors:** Mike Lonergan

**Author notes:** Corresponding author: Mike Lonergan, Division of Molecular and Clinical, Medicine, University of Dundee, Ninewells Hospital and Medical School, Dundee, DD1 9SY.

## Abstract

Re-examination of the large dataset collected and meta-analysed by Dr Chu and his colleagues contradicts their conclusions about the effects of separation distance on infection risk. Their conclusion was based on misunderstandings of the datasets. Each of these estimated risk relative to that incurred when touching infected individuals. Allowing for this suggests that the main advantage of social distancing, a perhaps 78% (95% CI 24, 92) reduction in risk of infection, occurs at distances below 1m. The data imply an 11% chance of further distances reducing the risk, with any effects likely to be small. However the limitations of the dataset do limit the strength of these conclusions.

## Introduction

Dr Chu and his SURGE co-authors have created a wide-ranging and informative systematic review and meta-analysis of studies on physical distancing, face masks, and eye protection to prevent person-to-person transmission of SARS-CoV-2 and COVID-19.[1] Their work is particularly important given the current discussions in many countries over the requirements necessary to prevent a resurgence of COVID-19 as lockdowns and other restrictions are eased. But we were surprised to find their figure 3A showing large risk-ratios at short distances and much smaller ones for larger minimum separation distances.

We would have expected the opposite, especially as they conclude that “protection was increased as distance was lengthened (change in relative risk [RR] 2·02 per m; p_interaction_=0·041; moderate certainty)”. We identified the cause of our confusion by examination of their figure 3B: their estimate of the risk at 2m effectively resulted from multiplying together the risks at 0, 1 and 2m. So where we had thought the estimated risk at 2m was relative to the risk from being in direct contact, the authors treated it as being relative to the risk at 1m separation. We reexamined the data SURGE used to determine which interpretation was correct.

### Data

Ki *et al*. [2] compared 49 people (staff and patients) who came within 2m of an index patient, with MERS in hospital in South Korea, to 29 who were more than 2m from the index patient but in the same room (their table 2). They identified 9 infections among these people (their table 1), as well as 2 other patients who didn’t enter the index patient’s room. Two of the 9 had unknown risk of contact, and were conservatively assigned to the over 2m group in SURGE’s analysis. Six of the seven who were assigned to the below 2m group were medical staff and included “Two ambulance paramedics … were also infected … They had briefly touched the index patient”, “1 radiologist … was diagnosed He had briefly touched the index patient” and “a visitor … who stood briefly (1–2 min) beside the bed of the index patient”.

**Table 1:**
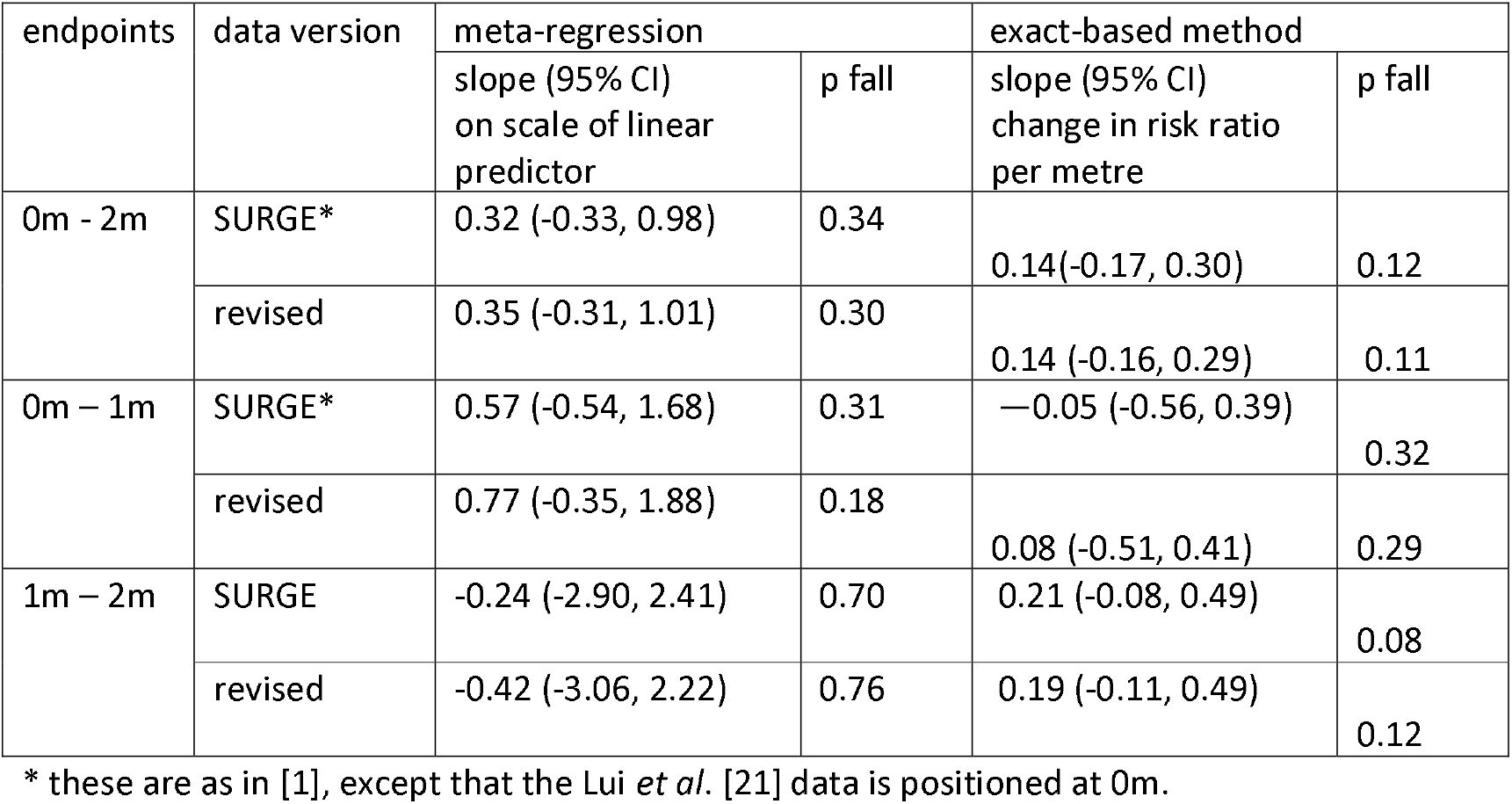
Effects of separation distance on the probability of infection. In each case p fall is the estimated probability that the longer distance provides a lower risk of infection than the shorter one. The estimates of slope are not directly comparable between the methods because they are measured on different scales.

Loeb *et al*. [3] compared 32 nurses who had entered the room of a patient, with SARS in a Canadian hospital, with 11 who hadn’t entered the room. SURGE treated this distinction as a 2m threshold. The list of procedures that were carried out includes intubation, manual dental care, and bathing. None of those seem consistent with avoiding contact and, additionally, the SURGE paper seems to double count those who caught the disease (Loeb table 1).

Yu *et al*. [4] looked at the acquisition of SARS by patients, within a ward in a Chinese hospital. SURGE treated being in a different bay as being beyond 2m. Yu’s paper says “Some inpatients in the ward were ambulatory” and shows a floor plan (their Figure 1) that suggests the beds were separated by less than 1m.

**Figure 1:**
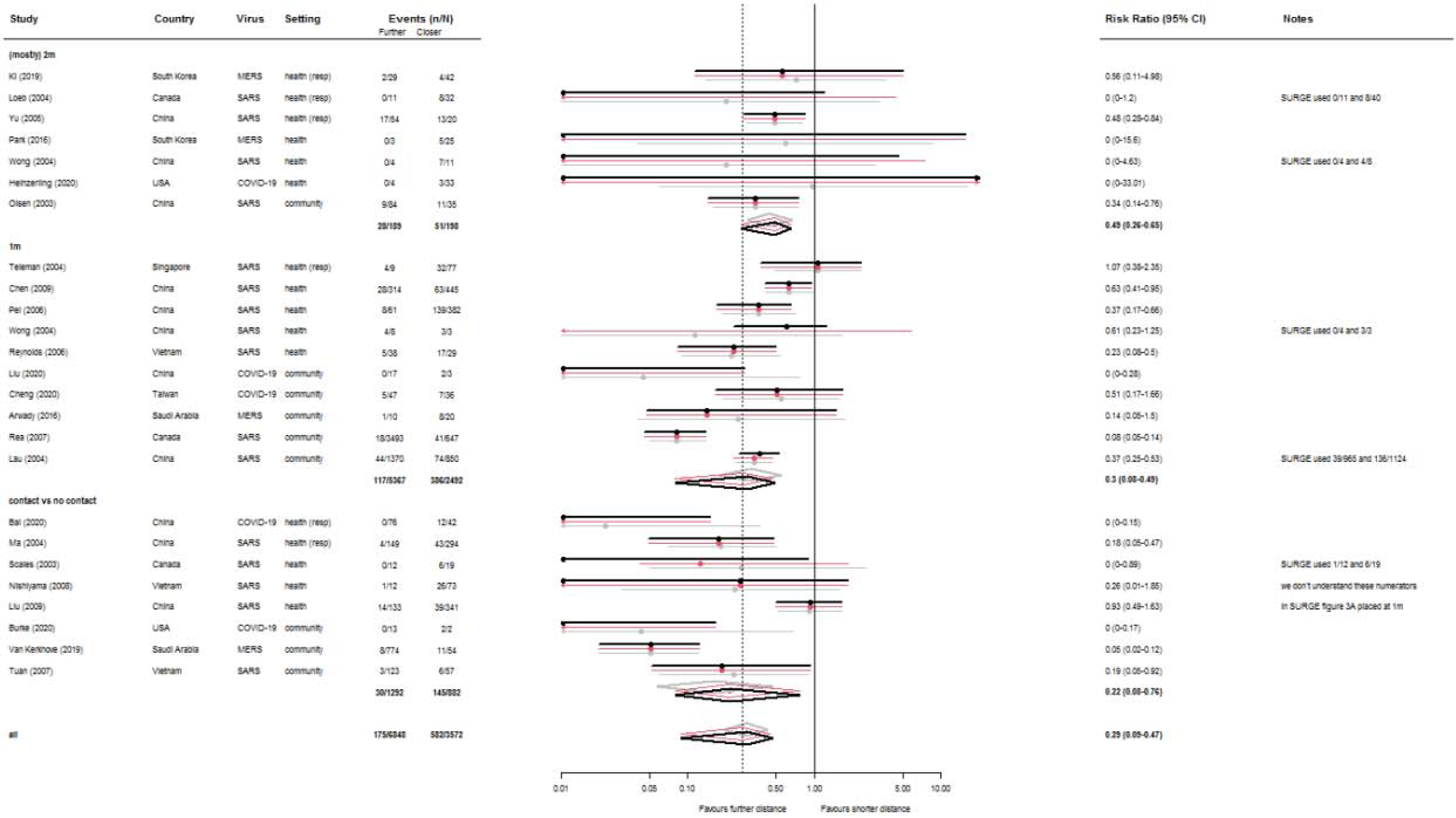
Forest plot of risk ratios estimated for each study, and groups of studies. Grey is estimates and 95% confidence intervals from the original SURGE[1] random effects meta-analysis. Red applies Tian’s exact method [33] to this data. Black applies the exact method to the revised data described in this paper. The Notes column describes the differences between the datasets, and the RR column gives numerical estimates for risk ratios in the revised dataset calculated by the exact method. The setting column indicates whether the data were collected from a health care setting (with resp indicating repirators were in use there) or in the wider community.

Park *et al*. [5] described 3 out of 5 of those, in their study within a South Korean hospital, infected at less than 2m as having “Touched the index case” (their Figure 2).

Heinzerling *et al*. [6] collected responses to questionnaires from 37 health care practitioners exposed a COVID-19 patient in a hospital in the USA. The three of them who became infected had been within 6’ (1.8m) of the patient (their Table 3), and were described as “one reported being present for a total of 3 hours while the patient was on BiPAP, and the other participated in BiPAP placement and intubation… The third staff member with COVID-19 reported close contact with the patient for a total of 2 hours”.

Olsen *et al*. [7] examined SARS transmission on board aircraft, and said “The WHO working definition of a contact on a flight is any passenger seated in the same row of seats or within two rows in front of or behind the index patient, or any flight attendant”. SURGE interpreted this split as a 1.5m threshold of separation distance.

The only study SURGE describe as investigating a threshold distance over 1m that that matches their interpretation is that by Wong *et al*. [8] It examined infections among medical students in a hospital in Hong Kong, and appears twice in both SURGE’s (their Figure 3A) and our forest plots (Figure 1) because they considered differences across 1m and 2m thresholds. None of the students physically examined the index patient. For our reanalysis we reassigned the individuals to match the definition used in the other studies.

A similar situation was visible at 1m. 69 of the 86 health care workers Teleman *et al*.[9] considered “touched patients”. SURGE interpreted often or every time answers to “Avoiding face to face while caring for patient” as a 1m separation, but 33 of the 749 reported “Performing tracheal intubations” (their Table 3). 18 of of the 29 individuals Reynolds *et al*. [10] report as coming with 1m of the index patient are also reported to have “Touched index patient” (their table 3). That could be considered as an additional line in the forest plot, though we haven’t made that change in order to maintain comparability with the SURGE paper.

The distinction Cheng *et al*. [11] make, which is interpreted as 1m by SURGE, is that a “contact was listed as a household contact if he/she lived in the same household with the index case, while those listed as family contacts were family members not living in the same household”. Arwady el al. [12] reported that 16 of the 30 subjects said they had kissed the infected individual. Rea *et al*. [13] gave “Providing direct care to a case, embracing a case, visiting a case” as examples of “Exposure for >30 min at a distance of <1 m” (their table 1). Similarly, 290 of those in the study by Lau *et al*. [14] reported “Sharing room and bed” with the index patient.

There were three papers that we couldn’t access [15-17], but we read through the rest [18-23]. Among these, the data from Scales *et al*. [19] was problematic: the two SURGE plots are inconsistent, but neither seems to match the original paper. That does show 6 of the 19 people who touched the patient becoming infected, but the 7 ^th^ infection wasn’t one of the other 12 people who entered the room. They were one of a much larger number “with no apparent direct exposure to the index patient”. The data from Liu *et al*. [21] is also wrongly positioned in SURGE’s Figure 3B: it was recorded as testing a separation distance of 0m, which is correctly shown in their figure 3A, but positioned at 1m in the plot of the meta-regression.

All the adjustments we made to the data are listed in Figure 1. Having confirmed that five of the studies [24-28] recorded no cases of disease transmission, we ignored them because the inclusion of non-event studies in meta-analyses is generally considered unwise. [29]

### Reanalysis

The papers the data came from seemed to fit with our initial assumption that all the estimated risks were relative to those of people who didn’t avoid direct physical contact with infected individuals. That seemed to leave three ways of explaining away the significant positive slope reported by SURGE: the data could be unsuitable for answering the question, there might be flaws in the analysis, or this could just be one of those 5% of times when a significant result occurs by chance.

One possibility was the sparsity of the data: standard meta-analysis techniques can fail when there are groups in which no events were observed. SURGE used “DerSimonian and Laird random-effects models”. That approach is widely used, but also known to struggle with small numbers and empty cells. [29] There are two specific difficulties: it uses large-sample approximations that can break down for rare events, and the convention of adding 0.5 to zero counts that enables computation can also bias the results.

We repeated SURGE’s unadjusted overall meta-analysis, using the metafor [30] library in R 4.0.0 [31]. We also used the exactmeta [32] R library to apply an alternative method [33] that does not require modifying the data. It estimates risk ratios as zero when the treatment group contains no events. We used the inverse-variance weighting within it, though the constant and fisher weightings produced very similar results.

SURGE’s estimate of the overall risk ratio from intervention, across all datasets was 0.30 (95% CI: 0.22, 0.44). Redoing the random effects meta-analysis to the original data but excluding the studies containing no events in the distant group gave 0.323 (95% CI: 0.214, 0.489). The exact method gave 0.273 (95% CI: 0.089, 0.451) for all the data and 0.271 (95% CI: 0.089, 0.459) excluding zeros. The main noticeable difference is that the exact method doesn’t impose symmetry, on the log-scale, on its confidence intervals. That allows their left-hand tails to stretch out towards zero.

We then carried out equivalent analyses for the contact *vs*. no contact studies (0m), those testing a 1m distancing, and the remainder. We consider that last analysis to represent 2m spacing because we believe 2m and 6’ thresholds to be effectively the same in this context, with the main actual difference being a matter of nationality. The one study with a 1.5m threshold is also included in this group; removing it made very little difference to the results.

Figure 1 shows the results for the data as we used it, and the differences from SURGE’s version. These differences largely cancel out, making minimal overall difference to the results. However, the existence of so many differences does indicate the difficulty and subjectivity of the task SURGE undertook.

### Effect of distance on risk ratios

It can be seen, in Figure 1, that the estimated risk ratios for the subgroups increase, i.e. approach 1, as the separation distance increases. When we repeated the meta-regression, we got the same result as SURGE: the risk ratio approaches 1 as the separation distance increases. However, the slope was no longer significant: simply transferring the Liu *et al*. 2009 data to its correct position increases the p-value from the 0.041 they reported (which our reanalysis estimated at 0.047) to 0.337. This large difference is because a high, precise value is moved from the centre of the data range, where it doesn’t affect the slope, to the lower end of the range, where it both raises the left-hand side of the line and increases the spread of the data there.

We fitted similar models, excluding either the 0m data or studies with thresholds greater than 1m, to give separate estimates of the changes over each metre. These gave non-significant results as well (Table 1). Rather than choose a baseline probability to convert the meta-regression confidence intervals into changes in risk, we generated scaled beta distributions that matched the means and the limits of the 95%confidence intervals of each subgroup’s estimate from the exact process. We drew 10000 times from each to generate distributions of values for each one. We then combined these into distributions of slopes and extracted estimates and confidence intervals for the rate of change of risk ratio with distance, plus associated p-values.

The two estimates of the rate at which infection risk changes with distance are not directly comparable, partly because they make different assumptions, but mainly because the meta-regression beta coefficient is on the scale of the model’s linear predictor while the exact-based estimator is directly a rate of change in risk-ratio. However, they both agree that these data suggest there is a fairly small chance, between 11% and 34%, that a 2m separation distance provides more protection from infection than simply avoiding direct physical contact (Table 1). The method based on the exact meta-analyses goes on to suggest that there is about a 2.5% chance that 2 metre separation reduces the chance of infection by 32% compared to simply avoiding direct contact. Comparisons over shorter distances are complicated by the long lower tail in the estimated risk ratio at 1m separation in the exact meta-analyses. These do not affect the meta-regression because of the assumptions of the random effects analysis. However it is notable that the exact-based method suggests that any benefit of increasing separation is likely to occur at below 1m distance, while the meta-regression suggests that it is more likely that 2m is better than 1m than it is that 1m separation is better than simply not touching.

The precisions of these estimates are poor, because of the limited amount of data on which they are based. So the exact-based method leaves open a 2.5% chance that moving out to 1m could produce a 49% reduction in risk, but suggests that the actual effect is likely to be closer to 8%.

## Discussion

It appears that SURGE’s identification of a strong and significant reduction in risk as separation distance increases out to 2m is based on a misinterpretation of the data within the original studies. These compared risk inside and beyond threshold distances, with many individuals in the closer in groups having physical contact with infected individual, even for large threshold distances. That means the risk ratios are all relative to those of touching, and should not be multiplied in the way Chu et al [1] seem to have done. The sparsity of the data, and some issues in its transcription had far less effect on the results.

This reanalysis suggests there may be some benefit in remaining 1m away from infected individuals, but that is likely to be small. It is much harder to find any support for advantages from greater separation distances here. It could be argued that it is obvious that increasing separation will reduce disease transmission, and therefore our results demonstrate the unsuitability of these data for investigating this issue. However, it seems unlikely that a better dataset will become available in the near future, so that would suggest science has little to contribute to decisions about social distancing.

It is often, rightly, been said that “extraordinary claims require extraordinary evidence”, but this situation may be a demonstration of the opposite: when we observe a result that fits our expectations we may be unlikely to question it sufficiently. Normally a positive regression coefficient, or line that rises to the right, indicates an increase. In this case it showed a ratio approaching unity. Additionally, the relationship appeared statistically significant. Clear answers are quite rare in this sort of science, so that would feel more a cause for celebration than a reason to worry. Given that this dataset is at the edge of what can support this sort of analysis, and the techniques were sophisticated and indirect, the confusion is understandable.

However, more important than why this reanalysis came to different conclusion that the original, is its practical implications. If simply not touching infected individuals is likely to reduce infection risk by more than half, and longer separation distances add little to this, then there may be little benefit from long-range social distancing.

These results could be taken as suggesting that the bulk of infection takes place through the direct transfer of material, rather than by an aerial route. If that is so, then, provided we refrain from coughing or spitting on each other, public health policy might be better concentrating more on limiting the touching of surfaces[34] than on keeping people apart. They seem to suggest that the observed benefits of social distancing have resulted more from reductions in contact with things others have recently touched than in our avoidance of their exhalations. Theyalso fit with the suggestion that the major benefit from wearing masks comes from reductions in touching of the face.

However, touching requires proximity. And, therefore, not touching infected individuals s likely to mean spending less time very close to them. So these data cannot determine how much of the benefit actually comes from the first decimetres of separation, rather than simply the lack of physical contact.

## Data Availability

The data is all tabulated in the manuscript.

